# Physician perspectives on low value care in Trinidad and Tobago- a cross sectional survey

**DOI:** 10.64898/2026.01.17.26344319

**Authors:** Loren De Freitas, Jillian Regobert, Saleem Varachhia

## Abstract

**Objective:** To evaluate the views of physicians in Trinidad and Tobago towards low value care

**Methods:** Physicians were surveyed using a cross sectional study design. An online questionnaire was disseminated through social media platforms using convenience sampling. A descriptive analysis was performed.

**Results:** Data from 218 physicians were analysed. Most participants worked in internal medicine (n=59, 27.1%) and the majority of participants were junior doctors (n=147, 67.4%). Most participants (n=97, 44.1%) said they rarely recommended low value care to patients while 51.8% (n=113) said their colleagues sometimes recommended low value care. Almost all participants (n=210, 97.1%) were interested in learning more about evidence-based recommendations that could address when a test/procedure is unnecessary. Reasons for ordering unnecessary tests included because it was an order from the senior doctor in the specialty or the admitting doctor requested the test (68.3%, n=149), inadequate information (61.5%, n=134), difficulty accessing prior medical records (126, n=57.8) and fear of litigation (57.3%, n=125). Strategies to reduce unnecessary care were training (92.2%, n=201), ease of access to external records (72.9%, n= 159), clinical pathways (64.2%, n= 140) and educational materials for patients (64.2%, n=140)

**Conclusion:** Low value care is an area of concern in Trinidad and Tobago. Identifying areas of overuse and developing targeted plans to reduce unnecessary care are important next steps.

## INTRODUCTION

Low value care, overuse, underuse, overdiagnosis and overtreatment are concepts that have been recognised as areas of concern in health systems globally. Low value care, sometimes known as unnecessary medical care, may be considered an intervention or clinical care that has little benefit for patients, or where the risk of harm exceeds potential benefit [1]. Overuse has been similarly defined [2] but Brownlee simply states overuse are services that are unnecessary [3]. The harms associated with overuse include psychological, physical harms and wasted resources in health systems [3]. International recommendations are attempting to reduce this low value care to improve the quality of care provided to patients. The European Commission Member States Expert Group on Health Systems Performance Assessment published a report on low value care in 2025 which explored definitions, indicators and de-implementation strategies for low value care [4]. One established organisation whose specific goal is to promote the reduction of low value care is the international Choosing Wisely campaign [5]. The core principle of Choosing Wisely is to promote conversations between physicians and patients to reduce unnecessary tests, treatment and procedures [5, 6].

The focus on low value care is particularly relevant now given the impact of the health sector on climate change with the global health sector contributing almost 5% of global carbon emissions [7]. Health sectors around the world are striving towards reducing their carbon footprints by encouraging environmentally sustainable clinical practices which includes reducing unnecessary tests and procedures [7]. Thus, prioritising reductions in low value care has a role to play in creating low carbon, climate resilient health systems. In the Caribbean, there is limited research on low value care. This study therefore aims to evaluate the views of physicians in Trinidad and Tobago towards low value care.

## METHODS

### Study setting and design

An online cross-sectional survey design was conducted, targeting physicians in Trinidad and Tobago. A 15-20 minute online questionnaire was used to collect data. The questionnaire mainly consisted of questions taken from the Unnecessary Tests and Procedures in the Health Care System, a questionnaire developed by the American Board of Internal Medicine Foundation (ABIM), as well as other studies that examined low value health care [8–10]. Permission was obtained from the ABIM to utilise their questionnaire.

### Target population and sampling

The target population were physicians licensed to practice medicine in Trinidad and Tobago, capable of understanding English and providing consent. A pragmatic approach using convenience sampling was used to access as many participants as possible. The calculated sample size was 351.

### Data collection

The questionnaire was piloted on a sample of 10 physicians with the purpose of assessing the understanding of the questions. The questionnaire was modified based on feedback. Data from the pilot were not used in the final analysis.

The main data collection occurred from August to October 2022. The survey was self-administered using an online platform, Google Forms and distributed using multiple social media platforms including Whatsapp and LinkedIn and via email through medical associations. Participation was voluntary, anonymous and confidential and no compensation was provided. An online participant information and consent sheet was included with the survey. Participants consented to participate in the survey prior to starting the survey. Ethical approval was granted by the Ministry of Health Ethics Committee of Trinidad and Tobago.

### Data analysis

Data were analysed using descriptive statistics using The Statistical Package for the Social Sciences, Version 23 (SPSS 23 for windows). Descriptive statistics, including frequencies and percentages for categorical variables and means and standard deviations for continuous variables, were used to summarize the characteristics of the study population and survey responses.

## RESULTS

### Demographics

A total of 220 responses were collected. Two responses were excluded because participants did not provide consent. Thus, data from 218 responses were analysed. Overall, 63.6% (n=138) of participants were female and the mean age was 32.7 years. The largest proportion of responses came from participants working in internal medicine (n=59, 27.1%), emergency medicine (n=52, 23.9%) and primary care (n=52, 23.9%). Further details are presented in Table 1.

**Table 1.**
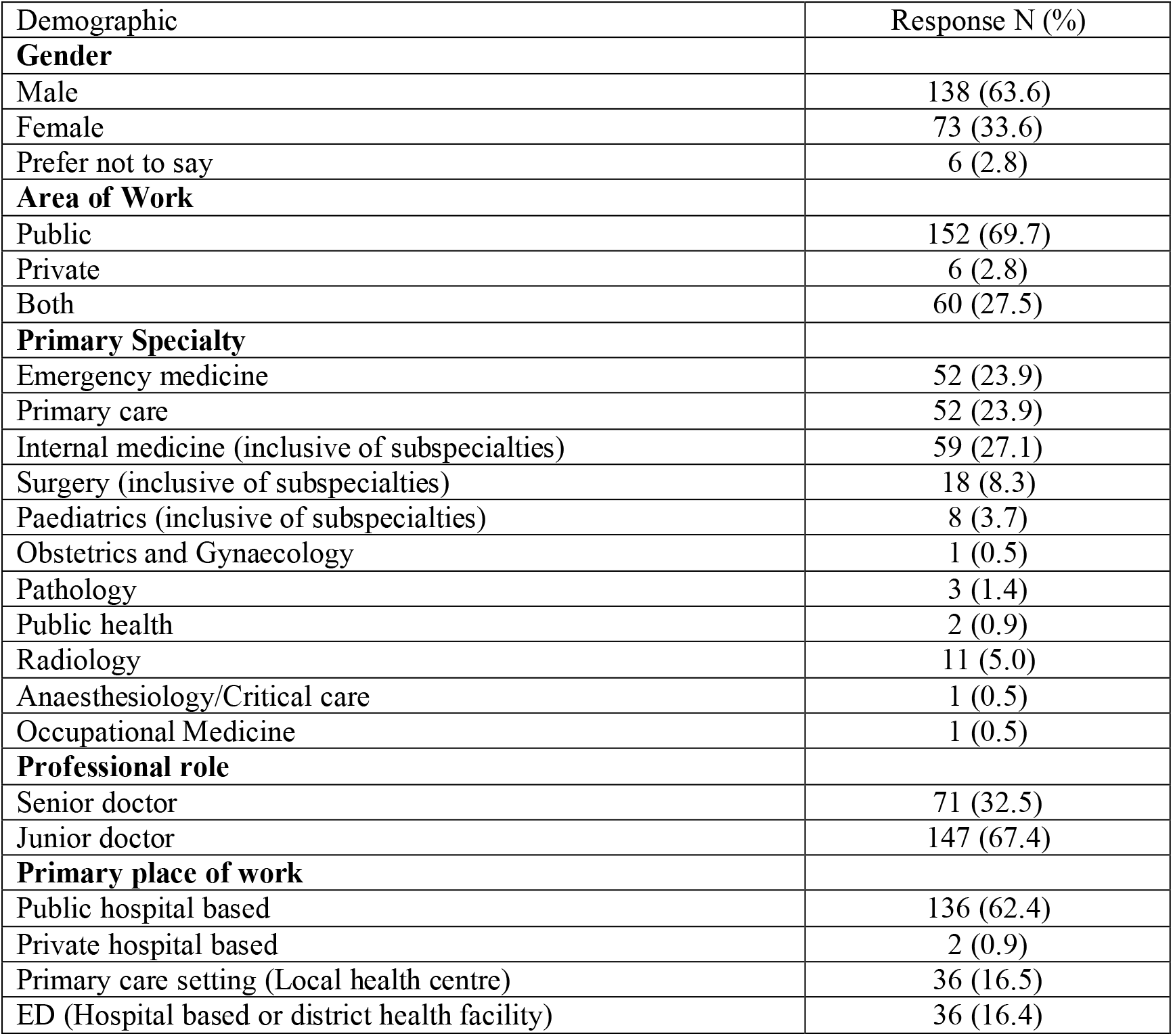
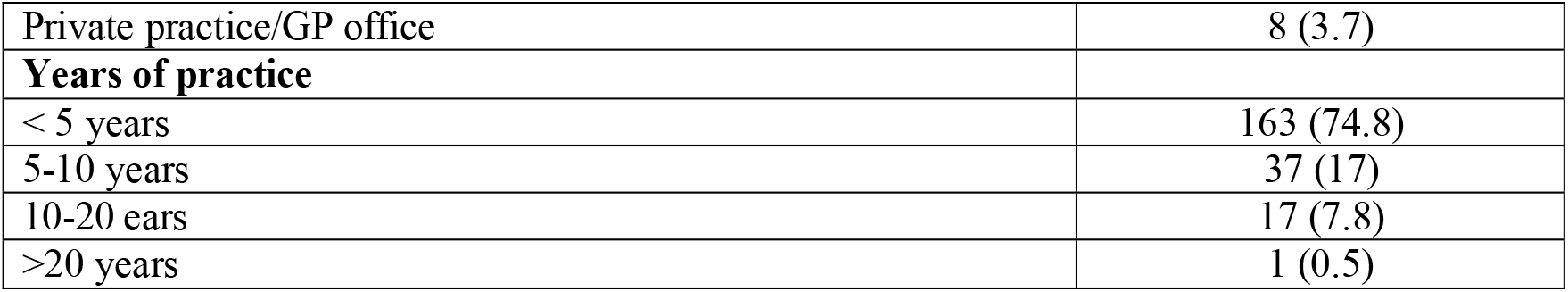
Physician Characteristics.

| Demographic | Response N (%) |
| --- | --- |
| **Gender** |  |
| Male | 138 (63.6) |
| Female | 73 (33.6) |
| Prefer not to say | 6 (2.8) |
| **Area of Work** |  |
| Public | 152 (69.7) |
| Private | 6 (2.8) |
| Both | 60 (27.5) |
| **Primary Specialty** |  |
| Emergency medicine | 52 (23.9) |
| Primary care | 52 (23.9) |
| Internal medicine (inclusive of subspecialties) | 59 (27.1) |
| Surgery (inclusive of subspecialties) | 18 (8.3) |
| Paediatrics (inclusive of subspecialties) | 8 (3.7) |
| Obstetrics and Gynaecology | 1 (0.5) |
| Pathology | 3 (1.4) |
| Public health | 2 (0.9) |
| Radiology | 11 (5.0) |
| Anaesthesiology/Critical care | 1 (0.5) |
| Occupational Medicine | 1 (0.5) |
| **Professional role** |  |
| Senior doctor | 71 (32.5) |
| Junior doctor | 147 (67.4) |
| **Primary place of work** |  |
| Public hospital based | 136 (62.4) |
| Private hospital based | 2 (0.9) |
| Primary care setting (Local health centre) | 36 (16.5) |
| ED (Hospital based or district health facility) | 36 (16.4) |
| Private practice/GP office | 8 (3.7) |
| **Years of practice** |  |
| < 5 years | 163 (74.8) |
| 5-10 years | 37 (17) |
| 10-20 ears | 17 (7.8) |
| >20 years | 1 (0.5) |

### Attitudes towards low value care

Less than 50% of participants (n=97, 44.1%) said they rarely recommended low value care to patients while 51.8% (n=113) said their colleagues sometimes recommended low value care. Most participants either rarely (n= 78, 35.8%) or sometimes (n=92, 41.8%) felt pressure from patients to order more tests and 31.7% (n=69) said patients asked for unnecessary tests/procedures a couple of times a month. When asked if the participant would order an unnecessary test because of the patient’s insistence 54.1% (n=118) stated they would refuse to order the test while 45.4% (n=99) said they would order the test but would give reasons why the test should not be ordered.

Most participants were somewhat comfortable discussing low value care with patients (n= 74, 33.9%) and colleagues (n=71, 32.6%). When asked how often participants talked to patients about why they should not have an unnecessary test, 50.5% (n=110) said they always talked to patients while 44% (n=96) said patients agreed to avoid an unnecessary test/procedure after talking to them. The majority of participants either rarely (n=78, 35.8%) or sometimes (n=74, 33.9%) talked to their colleagues about why they should not have ordered an unnecessary test/procedure. The majority of participants (n=210, 97.1%) were interested in learning more about evidence-based recommendations that could address when a test/procedure is unnecessary and 87.1% (n= 178) were not aware about the international Choosing Wisely campaign. Overall, more than 80% (n=182, 82.7%) of participants felt that up to 30% of care was unnecessary in their specialty and 41.3% (n=90) felt that the frequency of unnecessary tests/procedures were somewhat serious of a problem in the health system. Details are presented in Table 2.

**Table 2.** Attitudes towards low value care.

| **Variable** | **Response**<br>**N (%)** |
| --- | --- |
| **The care I recommend to my patients is of low value** |  |
| Never | 63 (28.9%) |
| Rarely | 97 (44.5%) |
| Sometimes | 51 (23.4%) |
| Very Often | 6 (2.8%) |
| Always | 1 (0.5%) |
| **The care my colleagues provide is low value** |  |
| Never | 17 (7.8%) |
| Rarely | 63 (28.9%) |
| Sometimes | 113 (51.8%) |
| Very Often | 25 (11.5) |
| Always | 0 (0.0%) |
| **I feel pressure from my patients to order more tests and procedures in my clinical practice** |  |
| Never | 27 (12.4%) |
| Rarely | 78 (35.8%) |
| Sometimes | 92 (42.2) |
| Very Often | 19 (8.7) |
| Always | 2 (0.9) |
| **I should be solely devoted to my individual patients' best interests, even if that is expensive** |  |
| Strongly disagree | 3 (1.4%) |
| Disagree | 28 (12.8%) |
| Neutral | 46(21.1%) |
| Agree | 111 (50.9%) |
| Strongly agree | 30 (13.8%) |
| **A patient came to you convinced he or she needed a specific test. You knew the test was unnecessary, but the patient was quite insistent.** |  |
| Order the test | 1 (0.5%) |
| Order the test, but give reasons why you advise against it | 99 (45.4%) |
| Refuse to order the test | 118 (54.1%) |
| **Comfort level in having discussions with *patients* about low value care** |  |
| Very uncomfortable | 18 (8.3%) |
| Somewhat uncomfortable | 33 (15.1%) |
| Neutral | 38 (17.4%) |
| Somewhat comfortable | 74 (33.9%) |
| Completely Comfortable | 55 (25.2%) |
| **Comfort level in having discussions with *colleagues* about low value care** |  |
| Very uncomfortable | 13 (6.0%) |
| Somewhat uncomfortable | 38 (17.4%) |
| Neutral | 32 (14.7%) |
| Somewhat comfortable | 71 (32.6%) |
| Completely Comfortable | 64 (29.4%) |
| **Frequency that patients *ask* for a test or procedure that you think is unnecessary in your practice** |  |
| Less than once a month | 63 (28.9%) |
| A Couple of times a month | 69 (31.7%) |
| Once a week | 34 (15.6%) |
| Several times a week | 40 (18.3) |
| Every day | 12 (5.5%) |
| **Frequency with which you *talk* to patients about why they should not have a test or procedure that you feel is unnecessary** |  |
| Never | 1 (0.5%) |
| Rarely | 3 (1.4%) |
| Sometimes | 19 (8.7%) |
| Very Often | 85 (39%) |
| Always | 110 (50.5) |
| **Frequency with which patients *agree* to avoid the unnecessary test or procedure when you to them about it** |  |
| Never | 5 (2.3%) |
| Rarely | 16 (7.3%) |
| Sometimes | 93 (42.7%) |
| Very Often | 96 (44%) |
| Always | 8 (3.7%) |
| **Frequency with which you *talk* to colleagues about why they should not have ordered the test or procedure that you felt was unnecessary** |  |
| Never | 30 (13.8%) |
| Rarely | 78 (35.8%) |
| Sometimes | 74 (33.9%) |
| Very Often | 31 (14.2%) |
| Always | 5 (2.3%) |
| **In your specialty, what percent of overall care do you think is unnecessary?** |  |
| Less than 10% | 74 (33.9%) |
| Between 10-20% | 66 (30.3%) |
| Between 21-30% | 42 (19.3%) |
| Between 31-40% | 21 (9.6%) |
| Greater than 40% | 15 (6.9%) |
| **Frequency of unnecessary tests and procedures in the health care system in T&T** |  |
| Not a problem at all | 2 (0.9%) |
| Not too serious a problem | 32 (14.7%) |
| Somewhat serious | 90 (41.3%) |
| Very serious problem | 81 (37.2%) |
| I don't know | 13 (6.0%) |
| **Requesting unnecessary tests negatively impacts/harms the environment** |  |
| Strongly disagree | 3 (1.4%) |
| Disagree | 14 (6.4%) |
| Neutral | 41 (18.8%) |
| Agree | 87 (39.9%) |
| Strongly agree | 60 (27.5%) |
| I don't know | 13 (6.0%) |
| **Interest in learning more about evidenced based recommendations to address unnecessary care** |  |
| Yes | 6 (2.9%) |
| No | 204 (97.1%) |
| **Awareness of Choosing Wisely campaign** |  |
| Not, not at all aware | 178 (81.7%) |
| Yes, somewhat aware | 35 (16.1%) |
| Yes, very aware | 5 (2.3%) |
| **Choosing wisely as a legitimate source of guidance** |  |
| No, not at all | 1 (0.5%) |
| Yes, somewhat | 26 (65%) |
| Yes, absolutely | 13 (32.9%) |

Most participants (n=87, 39.9%) agreed that unnecessary tests negatively impacted the environment. The majority of participants either agreed or strongly agreed that health professionals (n=158, 86.2%), health facilities (n=179, 82.1%) were in the best position to address the problem of unnecessary tests/procedures. Details of this are presented in Figure 1.

**Figure 1.**
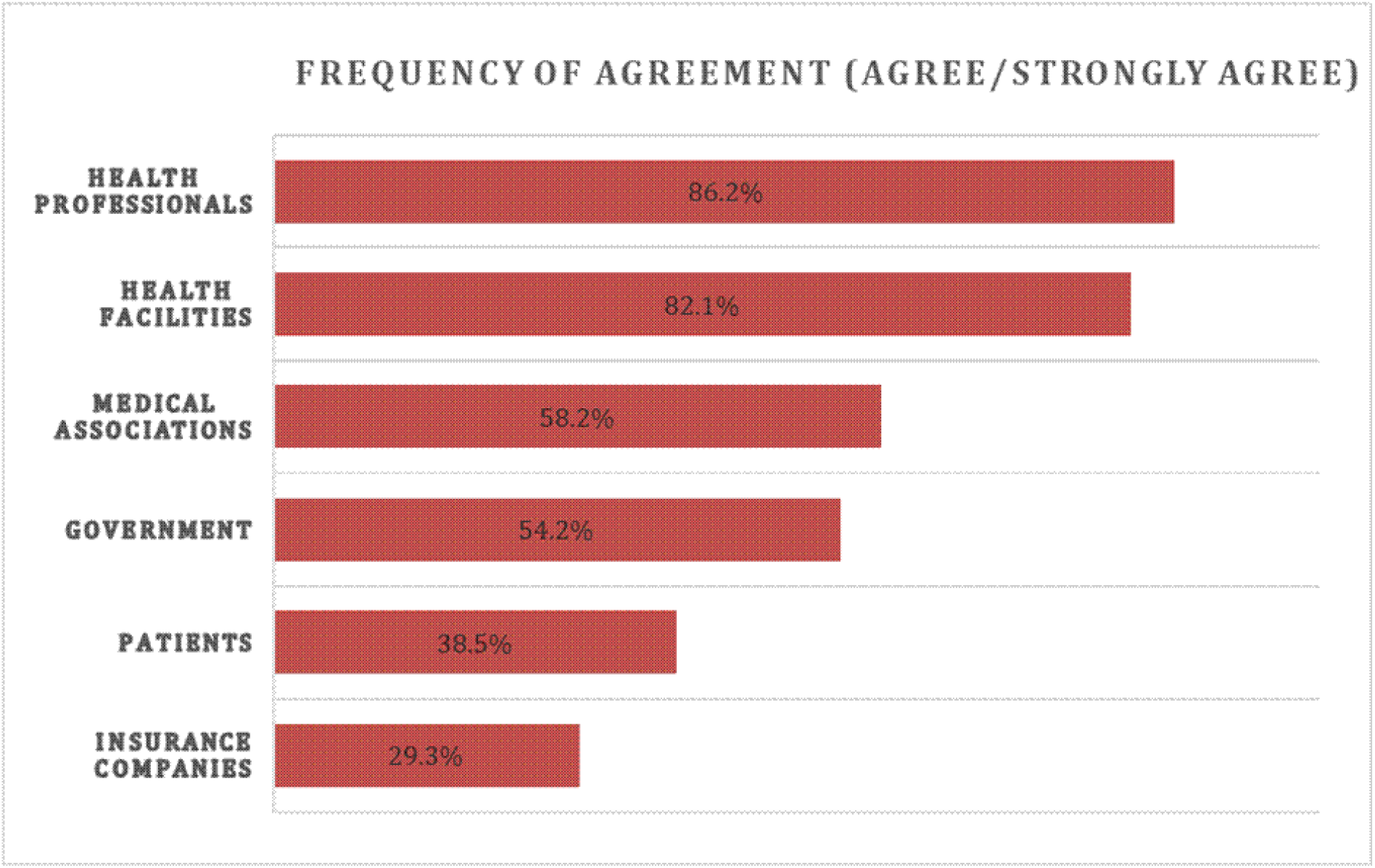
Participant views on who is responsible for addressing unnecessary care.

### Reasons for ordering unnecessary tests and strategies to reduce unnecessary testing

Physicians were asked about reasons for ordering unnecessary tests and strategies to reduce unnecessary care (Table 3). The top four reasons were: the test was requested by the senior doctor in the specialty or the admitting doctor (68.3%, n=149), inadequate information (61.5%, n=134), difficulty accessing prior medical records (126, n=57.8) and fear of litigation (57.3%, n=125). The top four strategies to reduce unnecessary care were training (92.2%, n=201), ease of access to external records (72.9%, n= 159), clinical pathways (64.2%, n= 140) and educational materials for patients (64.2%, n=140).

**Table 3.** Reasons for ordering and strategies to reduce unnecessary tests.

| **Reason for ordering unnecessary test** | **N (%)** | **Strategy to reduce unnecessary care** | **N (%)** |
| --- | --- | --- | --- |
| Senior doctor in your specialty or admitting doctor requests the test/procedure | 149 (68.3) | Training clinicians on evidence based guidelines | 201 (92.2) |
| Lack of adequate information/past medical history | 134 (61.5) | Easy access to outside health records | 159 (72.9) |
| Difficulty accessing prior medical records | 126 (57.8) | Creation of clinical pathways | 140 (64.2) |
| Fear of litigation/malpractice | 125 (57.3) | Patient educational materials | 140 (64.2) |
| Borderline clinical indications | 113 (51.8) | Clinician audit/feedback | 121 (55.5) |
| Want more information to reassure myself/just to be safe | 92 (42.2) | Having more time to discuss with patients | 100 (45.9) |
| Patients' preferences (ie the patient wants the test) | 69 (31.0) | Creation of organisational change frameworks | 86 (39.4) |
| Usual standard of practice in my workplace | 61 (28.0) | Listing prices when ordering tests | 83 (38.1) |
| Hassle of communicating with other physicians | 46 (21.1) | Increase base salaries and decrease fee-for service | 36 (16.5) |
| Fear of patients' satisfaction with me for not ordering the test/procedure | 37 (17.0) |  |  |
| Hospital or ED crowding | 25 (11.4) |  |  |
| Not enough time with patients | 24 (11.0) |  |  |
| Availability of tools to help efficiently share benefits and harms of low value care to patients | 20 (9.2) |  |  |
| Fee for service system of payment | 5 (2.3) |  |  |

## DISCUSSION

This study assessed the views of physicians in Trinidad and Tobago on low value care. Less than half of the participants felt they provided low value care and were comfortable discussing the topic with patients and almost all the participants wanted to learn more about evidence based recommendations on reducing low value care. Many participants also agreed that low value care had a negative impact on the environment. However, the majority of the participants were unaware about the international Choosing Wisely campaign. Key findings are discussed.

Most physicians did not think they practised low value care but many felt that their colleagues provided low value care and fewer physicians said they would speak with their colleagues who requested unnecessary tests compared to speaking with patients about the same topic. This is consistent with a US study which noted that fewer physicians were willing to speak with their colleagues about unnecessary care [11]. In our study, one potential reason could be a reflection of the participant group with the majority of participants being junior doctors who may not feel comfortable talking to their peers or their seniors if they disagree with a test.

The majority of physicians in our study considered the frequency of unnecessary care to be a problem in Trinidad and Tobago, with most reporting that up to 30% of care provided in their respective specialties was unnecessary. This is similar to another study where the majority of physicians felt that between 15-30% of care in the US was unnecessary [12] while a Saudi Arabian study of emergency physicians also reported rates of overuse from 30% to 60% depending on the clinical area [13] and is similar to global reports that indicate a quarter to a third of care is low value. These results suggest that unnecessary care is a problem across a range of settings and there is an opportunity to address waste in health care and maximise what is valuable to patients and physicians. Physicians also viewed health professionals and health facilities as those in the best position to address the issue of unnecessary care consistent with a Saudi Arabian study amongst emergency physicians [13].

When asked about the international Choosing Wisely campaign, most were unaware of the campaign but were interested in learning more about evidence to support reducing low value care. Since Choosing Wisely campaigns often partner with national medical associations to promote recommendations, it is necessary to increase awareness of the campaign and the importance and benefits of partnering with national associations in order to ensure successful campaigns. In 2024, Choosing Wisely Canada partnered with the Pan American Health Organisation to support and promote Choosing Wisely campaigns in Latin America and the Caribbean which indicates interest in the region towards reducing low value care [14].

Many participants agreed that unnecessary care contributed to environmental harm. Overutilisation of resources not only harms the health system by using scarce resources but has a larger impact of contributing to higher carbon footprints in health facilities. Reducing unnecessary care is a practical, feasible means of reducing the impact of health on climate change and creating environmentally sustainable health systems. Given the vulnerability of the Caribbean region towards climate change, this approach is particularly important to the development of sustainable, low carbon health systems.

Several studies have identified reasons for low value care [15, 16]. Reasons included a lack of time to discuss with patients, patient’s demands, fear of misdiagnosis, non-adherence to evidence based guidelines, and defensive medicine. Our study findings are consistent with the literature suggesting that low value care is similar across settings.

### Strengths and Limitations

This study utilised a cross sectional study design which only looks at perspectives at one point in time. The majority of participants were junior doctors who would have less experience and whose perspectives may differ from senior doctors. However, the results suggest this group needs support and guidance to practice evidence based medicine. Future studies should attempt to obtain broader participation and should utilise qualitative methods to provide greater insight into clinical decision making related to low value care.

## Conclusion

This study has provided insight into physician views on low value care in the Trinidad and Tobago health system. Future research and next steps would include understanding patient perspectives, assessing views on individual Choosing Wisely recommendations and implementing projects to specifically identify areas of overuse and develop targeted plans to reduce unnecessary care.

## Data Availability

All data produced in the present work are contained in the manuscript

